# Interpretable AI for beat-to-beat cardiac function assessment

**DOI:** 10.1101/19012419

**Authors:** David Ouyang, Bryan He, Amirata Ghorbani, Curt P. Langlotz, Paul A. Heidenreich, Robert A. Harrington, David H. Liang, Euan A. Ashley, James Y. Zou

## Abstract

Accurate assessment of cardiac function is crucial for diagnosing cardiovascular disease^1^, screening for cardiotoxicity^2,3^, and deciding clinical management in patients with critical illness^4^. However human assessment of cardiac function focuses on a limited sampling of cardiac cycles and has significant interobserver variability despite years of training^2,5,6^. To overcome this challenge, we present the first beat-to-beat deep learning algorithm that surpasses human expert performance in the critical tasks of segmenting the left ventricle, estimating ejection fraction, and assessing cardiomyopathy. Trained on echocardiogram videos, our model accurately segments the left ventricle with a Dice Similarity Coefficient of 0.92, predicts ejection fraction with mean absolute error of 4.1%, and reliably classifies heart failure with reduced ejection fraction (AUC of 0.97). Prospective evaluation with repeated human measurements confirms that our model has less variance than experts. By leveraging information across multiple cardiac cycles, our model can identify subtle changes in ejection fraction, is more reproducible than human evaluation, and lays the foundation for precise diagnosis of cardiovascular disease. As a new resource to promote further innovation, we also make publicly available one of the largest medical video dataset of over 10,000 annotated echocardiograms.

**Key Points:**

- Video based deep learning evaluation of cardiac ultrasound accurately identifies cardiomyopathy and predict ejection fraction, the most common metric of cardiac function.
- Using human tracings obtained during standard clinical workflow, deep learning semantic segmentation accurately labels the left ventricle frame-by-frame, including in frames without prior human annotation.
- Computational cardiac function analysis allows for beat-by-beat assessment of ejection fraction, which more accurately assesses cardiac function in patients with atrial fibrillation, arrhythmias, and heart rate variability.

## Introduction

Cardiac function is essential for maintaining normal systemic tissue perfusion with cardiac dysfunction manifesting as dyspnea, fatigue, exercise intolerance, fluid retention and mortality^1,3,4,6–9^. Impairment of cardiac function is labeled as “cardiomyopathy” or “heart failure” and is a leading cause of hospitalization in the United States and a growing global health issue^1,10,11^. A variety of methodologies have been used to quantify cardiac function and diagnose dysfunction. In particular, left ventricular ejection fraction (EF), the ratio of left ventricular end systolic and end diastolic volume, is one of the most important metrics of cardiac function, as it identifies patients who are eligible for life prolonging therapies^8,12^. However, there can be significant interobserver variability as well as inter-modality discordance based on methodology and modality^2,5,6,12–15^.

Human assessment of ejection fraction has variance in part due to common irregularity in the heart rate and the laborious nature of calculation limiting every beat quantification^5,6^. While the American Society of Echocardiography and the European Association of Cardiovascular Imaging guidelines recommend tracing and averaging up to 5 consecutive cardiac cycles if variation is identified, EF is often evaluated from tracings of only one representative beat or visually approximated if a tracing is deemed inaccurate^6^. This results in high variance and limited precision^6,16^ with interobserver variation ranging from 7.6% to 13.9%^5,13–16^. This variation is observed despite substantial training by those reading the EF. More precise evaluation of cardiac function is necessary, as even patients with borderline reduction in EF have been shown to have significantly increased morbidity and mortality^17–19^.

With rapid image acquisition, relatively low cost, and without ionizing radiation, echocardiography is the most widely used modality for cardiovascular imaging^20,21^. Being the most common first-line cardiovascular imaging modality, there is great interest in using deep learning techniques to determine ejection fraction^22–24^. Limitations in human interpretation, including laborious manual segmentation and inability to perform beat-to-beat quantification may be overcome by sophisticated automated approaches^6,25,26^. Recent advances in deep learning suggest that it can accurately and reproducibly identify human-identifiable phenotypes as well as characteristics unrecognized by human experts^25,27–29^.

To overcome current limitations of human assessment of the left ventricular ejection fraction, we propose EchoNet-Dynamic, an end-to-end deep learning approach for left ventricular labeling and ejection fraction estimation from input echocardiogram videos alone. We first perform frame-level semantic segmentation of the left ventricle with weak supervision from prior clinical expert labeling. The segmentations are then combined with the native echocardiogram videos as input for a 3-dimensional (3D) convolutional neural network (CNN) with residual connections. This approach provides interpretable tracings of the ventricle, which facilitate human assessment and downstream analysis, while leveraging the 3D CNN to fully capture spatiotemporal patterns in the video^6,30,31^.

## Results

EchoNet-Dynamic has three key components (Figure 1). First, we constructed a CNN model with atrous convolutions for frame-level semantic segmentation of the left ventricle. Atrous convolutions has been previously shown to perform well on non-medical imaging datasets^30^. The standard human clinical workflow for estimating ejection fraction requires manual segmentation of the left ventricle during end-systole and end-diastole. We generalize these labels in a weak supervision approach with atrous convolutions to generate frame-level semantic segmentation throughout the cardiac cycle in a 1:1 pairing with the original video. This automatic segmentation improves the robustness of our model and make it more interpretable to clinicians.

**Figure 1.**
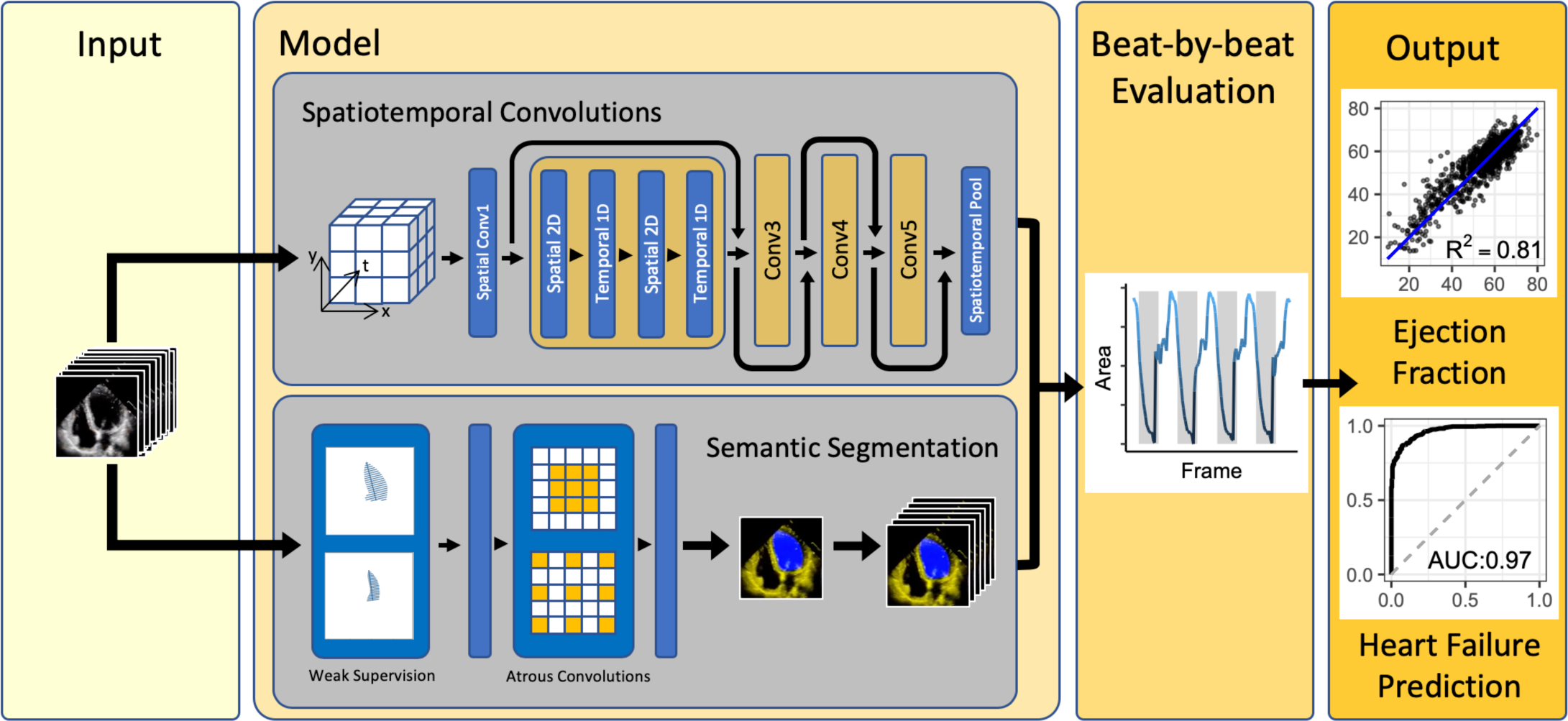
EchoNet-Dynamic workflow. For each patient, EchoNet-Dynamic uses standard apical-4-chamber view echocardiogram video as input. The model first predicts ejection fraction for each cardiac cycle using 3D spatiotemporal convolutions with residual connections and generates frame-level semantic segmentations of the left ventricle using weak supervision from expert human tracings. These outputs are combined to create beat-by-beat predictions of ejection fraction and to predict the presence of heart failure with reduced ejection fraction.

Second, we trained a CNN model with residual connections and 3D spatiotemporal convolutions across frames to predict ejection fraction. Unlike prior 3D CNN architectures for medical imaging machine learning, our approach integrates spatial as well as temporal information with temporal variation across frames as the third dimension in our network convolutions^25,31,32^. Spatiotemporal convolutions, which incorporate spatial information in two dimensions as well as temporal information in the third dimension has been previously used in non-medical video classification tasks^31,32^, however has not been previously attempted on medical imaging given the relative scarcity of video medical imaging datasets nor used for regression tasks instead of classification tasks.

Finally, we make video-level predictions of ejection fraction for beat-to-beat estimation of cardiac function. Each echocardiogram video typically includes multiple cardiac cycles, or beats, with each cycle being sufficient to produce a point estimate for ejection fraction. Given variance in cardiac function caused by changes in loading conditions as well as heart rate in a variety of cardiac conditions, it is recommended to perform ejection fraction estimation in up to 5 cardiac cycles, however this is not always done in clinical practice given the tedious and laborious nature of the calculation^6,16^. Our model identifies each cardiac cycle, generates a subsampled video-clip of 32 frames, and averages clip-level estimates of EF as a form of test-time augmentation.Details of the model and hyperparameter search is further described in Methods, Supplementary Table 1, and Supplementary Figure 1.

EchoNet-Dynamic was developed using 10,030 apical-4-chamber echocardiograms obtained through the course of routine clinical practice at Stanford hospital. Each echocardiogram video corresponds to a unique patient during a unique visit and is representative of the variation in patient characteristics and image acquisition at the hospital. Table 1 contains the summary statistics of the patient population. These randomly selected patients have a range of ejection fractions representative of the patient population going through the echocardiography lab and the echocardiogram videos were split 7,465, 1,277, and 1,288 patients respectively for the training, validation, and test sets.

**Table 1.**
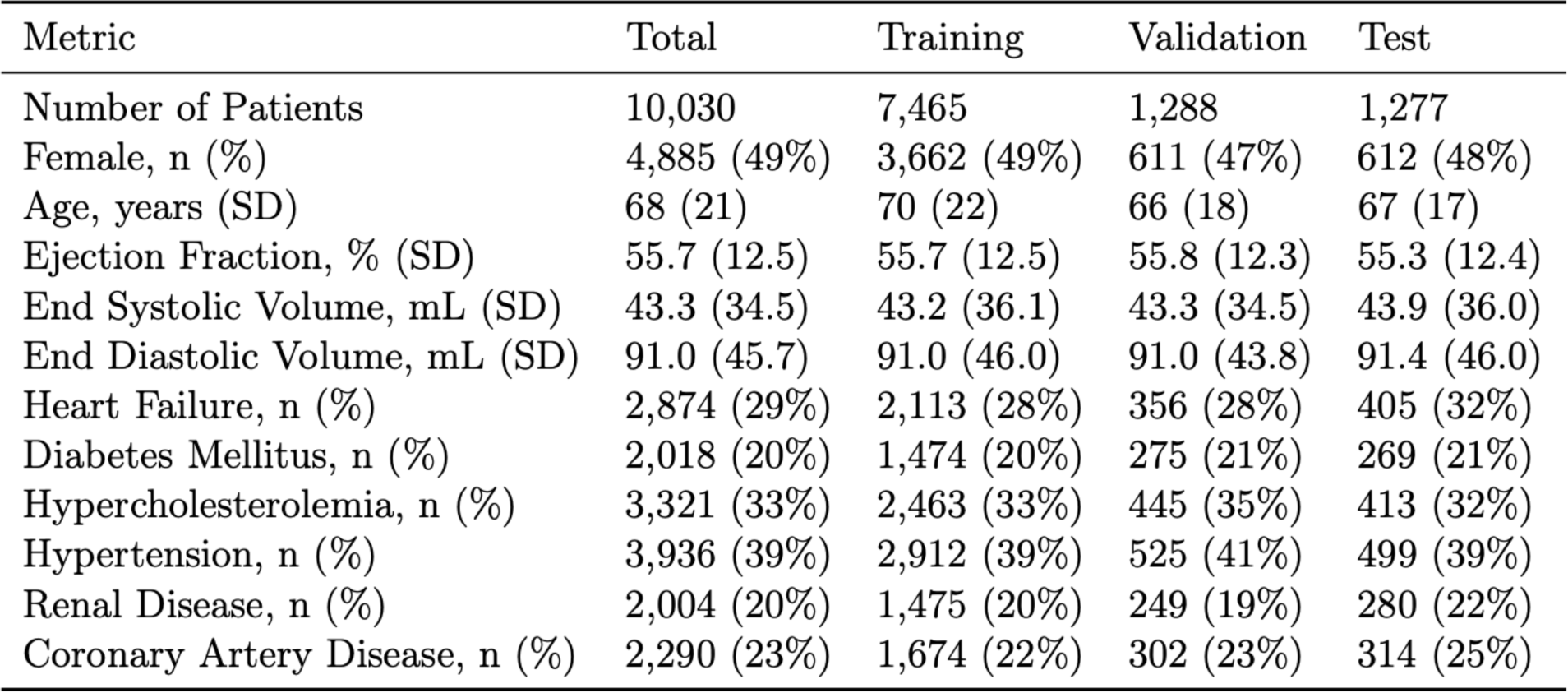
Summary statistics of patients in the dataset. Data obtained from visits to Stanford Hospital between 2016 and 2018.

We worked with Stanford University and Hospitals to release our full dataset of 10,030 de-identified echocardiogram videos as a resource for the medical machine learning community for future comparison and validation of deep learning models. To the best of our knowledge, this is the largest labeled medical video dataset to be made publicly available and first large release of echocardiogram data with matched labels of human expert tracings, volume estimates, and left ventricular ejection fraction calculation. We expect this dataset to greatly facilitate new echocardiogram and medical video based machine learning work.

In a test dataset not previously seen during model training, model performance on individual subsampled video clips of approximately 1 second had a mean absolute error of 4.2% (95% CI 4.0% - 4.3%), root mean squared error of 5.6% (5.7% - 5.8%) and R^2^ of 0.79 (95% CI 0.77 - 0.81) compared with the clinician report (Figure 2). Given that the model is agnostic to cardiac rhythm disturbances, including premature atrial contractions, premature ventricular contractions, and atrial fibrillation, we perform test time augmentation with beat-to-beat evaluation of ejection fraction. The final model with augmentation has improved performance with mean absolute error of 4.1%, root mean squared error of 5.3% and R^2^ of 0.81 (95% CI 0.78 - 0.82), which are within the range of typical measurement variation between different clinicians. We compared EchoNet-Dynamic’s performance with that of several additional deep learning models that we trained on this dataset, and EchoNet-Dynamic is consistently more accurate, suggesting the power of its specific architecture (Supplementary Table 1).

**Figure 2.**
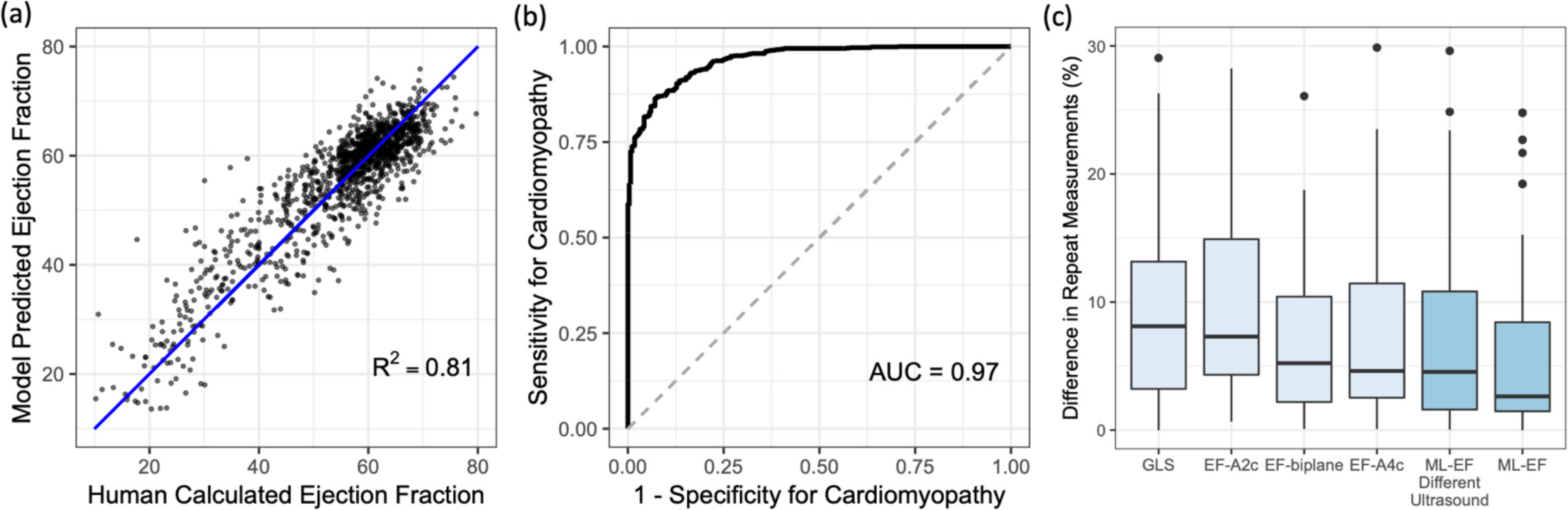
Model Performance. (a) EchoNet-Dynamic’s predicted ejection fraction vs. reported ejection fraction. (b) Receiver operating characteristic curve for diagnosis of heart failure with reduced ejection fraction. (c) Variance of metrics of cardiac function on repeat measurement. The first four boxplots corresponds to variation by clinicians using different techniques, and the last two boxplots corresponds EchoNet-Dynamic’s variance on input images from standard ultrasound machines and an ultrasound machine not previously seen by the model.

EchoNet-Dynamic was compared against human measurements on 55 patients prospectively evaluated by two different sonographers on the same day. Each patient was independently evaluated for global longitudinal strain (GLS) and ejection fraction by multiple methods as well as our model for comparison (Figure 2D). EchoNet-Dynamic assessment of cardiac function had the least variance on repeat testing (median difference of 2.6%, SD=6.4) compared to EF obtained by Simpson’s biplane method (median difference of 5.2%, SD=6.9, p < 0.001 for non-inferiority), EF from Simspon’s monoplane method (median difference of 4.6%, SD=7.3 p < 0.001 for non-inferiority), or GLS (median difference of 8.1%, SD=7.4% p < 0.001 for non-inferiority). Of the initial 55 patients, 49 patients were also assessed with a different ultrasound system never seen during model training and EchoNet-Dynamic assessment had similar variance (median difference of 4.5%, SD=7.0, p < 0.001 for non-inferiority for all comparisons with human measurements).

EchoNet-Dynamic automatically generates segmentations of the left ventricle, which enables clinicians to better understand how it makes predictions. The segmentation is also useful because this provides a relevant point for human interjection in the workflow and physician oversight of the model in clinical practice. For the semantic segmentation task, the labels were 20,060 frame-level labels of the left ventricle obtained during the course of standard human clinical workflow during which expert human sonographers and echocardiographers manually label of the left ventricle during end-systole and end-diastole. Given the average video contains 2 labeled frames but 176 total frames, these weak labels were used to generate frame-level segmentations for the entire video (Figure 3). On the test dataset, the Dice Similarity Coefficient (DSC) for the end systolic tracing was 0.903 (95% CI 0.901 – 0.906) and the DSC for the end diastolic tracing was 0.927 (95% CI 0.925 – 0.928). Despite being a frame-level, there was significant concordance in performance of end-systolic and end-diastolic semantic segmentation (Supplementary Figure 2). Example videos with semantic segmentation can be found in the Online Supplement.

**Figure 3.**
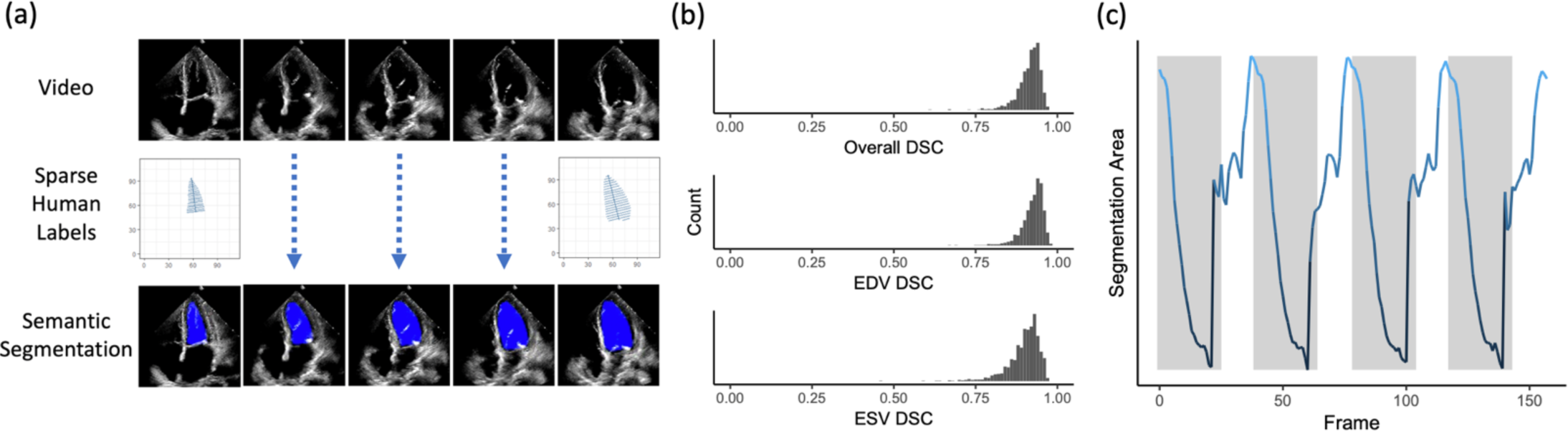
Semantic Segmentation Performance. (a) Weak supervision with human expert tracings of the left ventricle at end-systole and end-diastole is used to train a semantic segmentation model with input video frames throughout the cardiac cycle. (b) Dice Similarity Coefficient (DSC) was calculated for each ESV/EDV frame. (c) The area of the left ventricle segmentation was used to identify heart rate and bin clips for beat-to-beat evaluation of ejection fraction.

Variation in frame-to-frame model interpretation was seen in echocardiogram videos with arrhythmias and ectopy (Figure 4). In addition to correlation with irregularity in intervals between ventricular contractions, these videos were independently reviewed by clinical cardiologists and found to have atrial fibrillation, premature atrial contractions, and premature ventricular contractions. This highlights why it is important that EchoNet-Dynamic segments the ventricle and estimates the EF for each every beat in the video and then aggregates across the beats. In particular, by aggregating across multiple beats, EchoNet-Dynamic significantly reduces variation compared to the common clinical practice of estimating EF from a single beat (Figure 4d).

**Figure 4.**
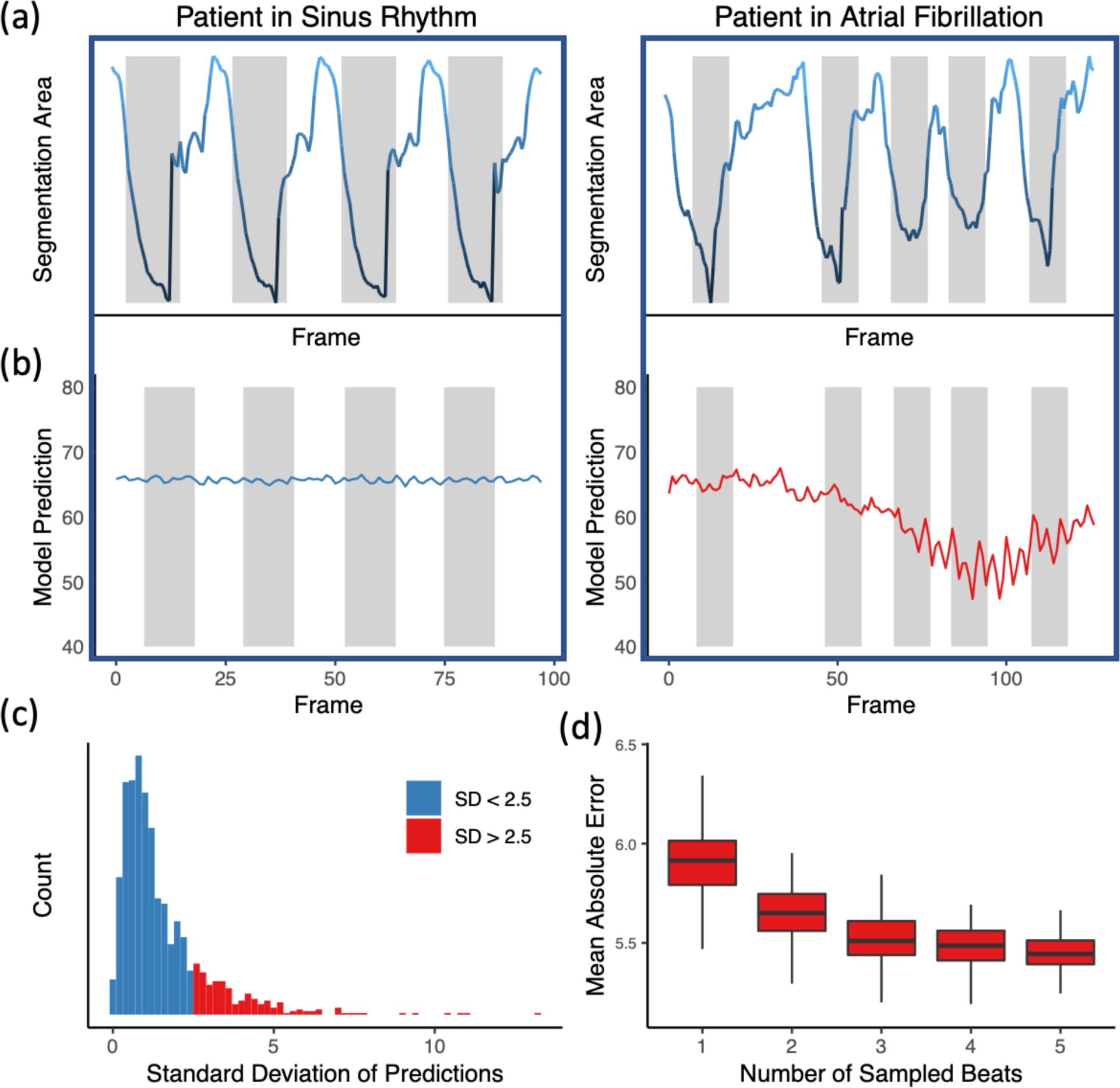
Beat-to-beat evaluation of ejection fraction. (a) Atrial fibrillation and arrhythmias can be identified by significant variation in intervals between ventricular contractions. (b) Significant variation in left ventricle segmentation area was associated with higher variance in EF prediction. (c) Histogram of standard deviation of beat-to-beat evaluation of EF across all the test videos. (d) Assessing the effect of beat-to-beat based on the number of sampled beats averaged for prediction. Each boxplot represents 100 random samples of a certain number of beats and comparison with reported ejection fraction.

## Discussion

EchoNet-Dynamic is a new video deep learning technique that achieves state-of-the-art assessment of cardiac function. It uses expert human tracings for weak supervision of left ventricular segmentation, 3D spatiotemporal convolutions on video data, and beat-to-beat cumulative evaluation of EF across the entire video. EchoNet-Dynamic’s performance in assessing EF is substantially better than prior deep learning attempts to assess EF^22^, and our model’s variance is less than human expert measurements of cardiac function. EchoNet-Dynamic could potentially aid clinicians with more precise and reproducible assessment of cardiac function and detect subclinical change in ejection fraction beyond the precision of human readers. Furthermore, we release the largest annotated medical video dataset, which will stimulate future work on machine learning for cardiology.

EchoNet-Dynamic diverged the most from human estimation of EF in videos with arrhythmias and variation in heart rate. This variation is a feature of comparing EchoNet-Dynamic’s beat-to-beat evaluation of EF across the video with our human evaluations of only one ‘representative’ beat. Choosing the representative beat can be subjective, contribute to human intra-observer variability, and less optimal compared to the guideline recommendation of averaging 5 consecutive beats. This workflow, is rarely done, in part due to the laborious and time intensive nature of the human tracing task. EchoNet-Dynamic greatly decreases the labor for cardiac function assessment with automating of the segmentation task and provide the opportunity for more frequent, rapid evaluation of cardiac function. Our end-to-end approach generates beat and clip level predictions of ejection fraction as well as segmentation of the left ventricle throughout the cardiac cycle for visual interpretation of the modeling results. In settings such as between chemotherapy sessions, after a heart transplant, and with the initiation of heart failure therapy, early detection of change in cardiac function significantly affect clinical care ^2,3^.

Future studies will be required to ensure clinical applicability as well as generalizability in different clinical scenarios and health systems. With rapid expansion in the use of point of care ultrasound for evaluation of cardiac function by non-cardiologists, we aim to explore the feasibility and generalizability of our model with input videos are variable quality and acquisition expertise. Correlating the model performance with improved clinical outcomes and health system costs will also be required to determine potential impact. In addition to its application assessing left ventricular ejection fraction, the deep learning techniques applied in this study have considerable relevance to other types of medical video imaging data with temporal information, including cardiac magnetic resonance imaging, as well as other functional assessments using echocardiogram videos.

These results represent an important step towards automated evaluation of cardiac function from echocardiogram videos through deep learning. EchoNet-Dynamic could augment current methods with improved precision, accuracy, and allow earlier detection of subclinical cardiac dysfunction, and the underlying dataset can be used to advance future work in deep learning for medical video imaging datasets and lay the groundwork for further applications of medical deep learning.

## Data Availability

The data is publicly available with a noncommerical data use agreement.

https://douyang.github.io/EchoNetDynamic/

## Acknowledgements

This work is supported by the Stanford Translational Research and Applied Medicine pilot grant, Stanford Cardiovascular Institute pilot grant, and a Stanford Artificial Intelligence in Imaging and Medicine Center seed grant. D.O. is supported by the American College of Cardiology Foundation / Merck Research Fellowship. B.H. is supported by the NSF Graduate Research Fellowship. A.G. is supported by the Stanford-Robert Bosch Graduate Fellowship in Science and Engineering. J.Z. is supported NSF CCF 1763191, NIH R21 MD012867-01, NIH P30AG059307 and by a Chan-Zuckerberg Biohub Fellowship.

## Author Contributions

DO retrieved and quality controlled all videos and merged electronic medical record data. DO, BH, AG, JYZ developed and trained the deep learning algorithms, performed statistical tests, and created all the figures. DO, CPL, PAH, RAH coordinated public release of the deidentified echocardiogram dataset. DO, PAH, DHL, EAA performed clinical evaluation of model performance. DO, BH, EAA, JYZ wrote the manuscript with all authors.

## Online Methods

### Data Curation

A standard full resting echocardiogram study consists of a series of 50-100 videos and still images visualizing the heart from different angles, locations, and image acquisition techniques (2D images, tissue Doppler images, color Doppler images, and others). In this dataset, one apical-4-chamber 2D gray-scale video is extracted from each study. Each video represents a unique individual as the dataset contains 10,025 echocardiography videos from 10,025 unique individuals who underwent echocardiography between 2016 and 2018 as part of clinical care at Stanford University Hospital. Images were acquired by skilled sonographers using iE33, Sonos, Acuson SC2000, Epiq 5G, or Epiq 7C ultrasound machines and processed images were stored in Philips Xcelera picture archiving and communication system. Video views were identified through implicit knowledge of view classification in the clinical database by identifying images and videos labeled with measurements done in the corresponding view.

The apical-4-chamber view video was identified by extracting the Digital Imaging and Communications In Medicine (DICOM) file linked to measurements of ventricular volume used to calculate the ejection fraction. Videos were spot checked for quality control, confirm view classification, and exclude videos with color Doppler. Each subsequent video was cropped and masked to remove text, ECG and respirometer information, and other information outside of the scanning sector. The resulting square images were either 600×600 or 768×768 pixels depending on the ultrasound machine and downsampled by cubic interpolation using OpenCV into standardized 112×112 pixel videos.

This research was approved by the Stanford University Institutional Review Board and data privacy review through a standardized workflow by the Center for Artificial Intelligence in Medicine and Imaging (AIMI) and the University Privacy Office. In addition to masking of text, ECG information, and extra data outside of the scanning sector in the video files as described below, each DICOM file’s pixel data was parsed out and saved as an AVI file to prevent any leakage of identifying information through public or private DICOM tags. Each video was subsequently manually reviewed by an employee of the Stanford Hospital with familiarity with imaging data to confirm the absence of any identifying information.

### Prospective Clinical Validation

Prospective validation was performed by two senior sonographers with advanced cardiac certification and greater than 15 years experience each. For each patient, measurements of cardiac function was independently acquired, measured, and assessed by each sonographer on the same day. Every patient was scanned using Epiq 7C ultrasound machines, the standard instrument in the Stanford Echocardiography Lab, and a subset of patients were also rescanned by the same two sonographers using a GE Vivid 95E ultrasound machine. Tracing and measurement was done on a dedicated workstation after image acquisition. For comparison, the independently acquired apical-4-chamber videos were fed into the model and the variance in measurements assessed.

### EchoNet-Dynamic development and training

Model building and training was done in Python on the PyTorch deep learning library. Semantic segmentation was performed using the Deeplabv3 architecture^30^. The segmentation model had a base architecture of 50-layer residual net and minimized a pixel level binary cross entropy loss. The model was initialized with random weights, and was trained using a stochastic gradient descent optimizer with a learning rate of 0.00001, momentum of 0.9, and batch size of 20 for 50 epochs. Our model with spatiotemporal convolutions was initialized with pretrained weights from the Kinetics-400 dataset^33^. We tested three model architectures with variable integration of temporal convolutions and ultimately chose decomposed R2+1D spatiotemporal convolutions as the model with the best performance^31,32^. The models were trained to minimize the squared loss between the prediction and true ejection fraction using a stochastic gradient descent optimizer with an initial learning rate of 0.0001, momentum of 0.9, and batch size of 16 for 45 epochs. The learning rate was decayed by a factor of 0.1 every 15 epochs was used during model training. During training, clips of 32 frames were generated by sampled every other frame. To augment the clips, all frames were padded with 12 pixels, and a random crop of the 112×112 pixel size was taken. For all models, the weights from the epoch with the lowest validation loss was selected for final testing.

### Test Time Augmentation with Beat-by-Beat Assessment

There can be variation in the ejection fraction, end systolic volume, and end diastolic volumes during atrial fibrillation, and in the setting of premature atrial contractions, premature ventricular contractions, and other sources of ectopy. The clinical convention is to identify at least one representative cardiac cycle and use this representative cardiac cycle to perform measurements, although an average of the measurements of up to five cardiac cycles is recommended when there is significant ectopy or variation. For this reason, our final model used test time augmentation by providing individual estimates for each ventricular beat throughout the entire video and outputs the average prediction as the final model prediction. We use the segmentation model to identify the area of the left ventricle and threshold-based processing to identify ventricular contractions during each cardiac cycle. For beat, a subsampled clip centered around the ventricular contraction was obtained and used to produce a beat-by-beat estimate of EF. The mean ejection fraction of all ventricular contractions in the video was used as the final model prediction.

### Statistical Analysis

Confidence intervals were computed using 10,000 bootstrapped samples and obtaining 95 percentile ranges for each prediction. Chi-squared test and Student’s t-test were used for statistical comparisons.

### Data Availability

This data introduces the EchoNet-Dynamic Dataset, a publicly available dataset of deidentified echocardiogram videos, publicly available at: https://douyang.github.io/EchoNetDynamic/

### Code Availability

The code is available at: https://github.com/douyang/EchoNetDynamic/

**Supplementary Figure 1:**
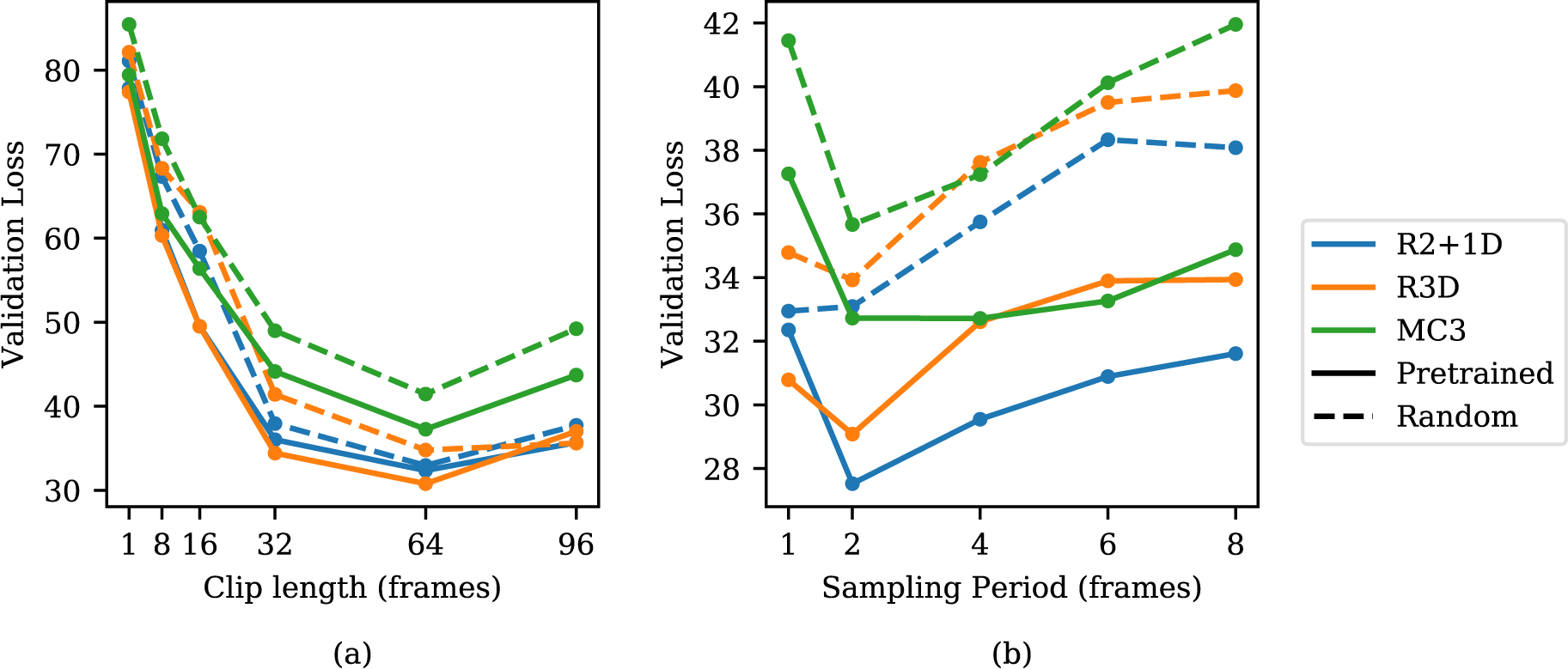
Hyperparameter search for 3D Spatiotemporal Convolutions on video dataset to predict ejection fraction. Model architecture (R2+1D, R3D, and MC3), initialization (Kinetics-400 pretrained weights with solid line and random initial weights with dotted line), clip length (1, 8, 16, 32, 64, and 96), and sampling period (1, 2, 4, 8) were considered. (a) When varying clip lengths, performance is best at 64 frames (corresponding to 1.28 seconds), and starting from pretrained weights improves performance slightly across all models. (b) Varying sampling period with a length to approximately correspond to 64 frames prior to subsampling. Performance is best at a sampling period of 2.

**Supplementary Figure 2:**
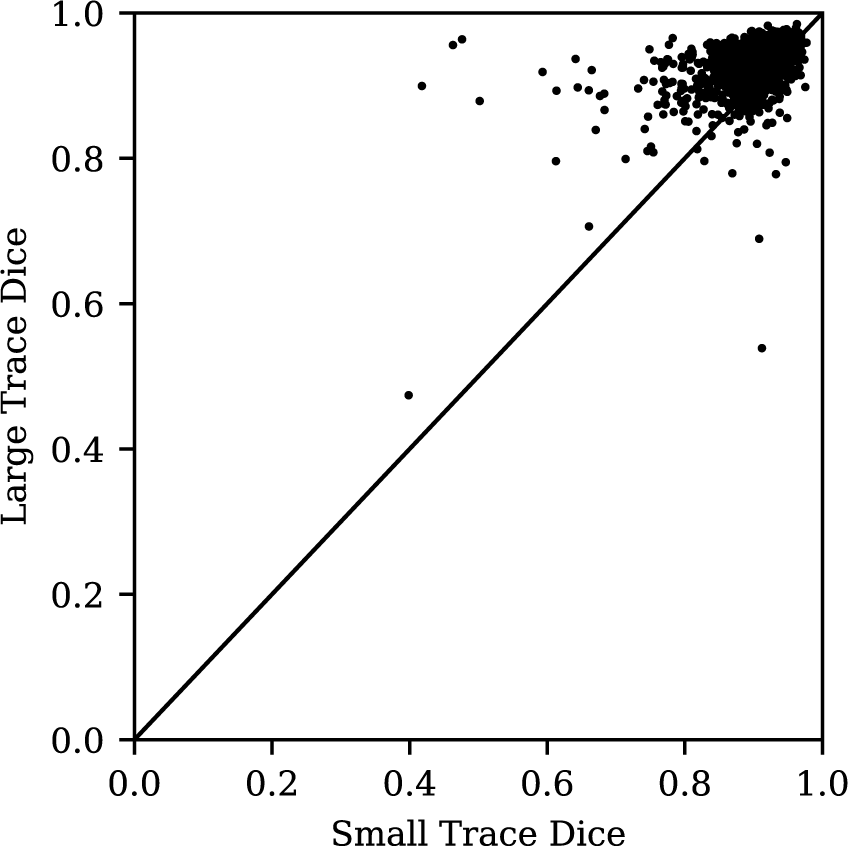
Relationship between end systolic tracing Dice Similarity Coefficient and end diastolic tracing Dice Similarity Coefficient.

**Supplementary Table 1:** Model performance compared to three alternative deep learning models in assessing cardiac function.

| Model | Evaluation | Sampling Period | MAE | RMSE | $R^2$ |
| --- | --- | --- | --- | --- | --- |
| EchoNet-Dynamic | Beat-by-beat | 1 in 2 | 4.05 | 5.32 | 0.81 |
| R2+1D | 32 frame sample | 1 in 2 | 4.22 | 5.56 | 0.79 |
| R3D | 32 frame sample | 1 in 2 | 4.21 | 5.62 | 0.79 |
| MC3 | 32 frame sample | 1 in 2 | 4.54 | 5.97 | 0.77 |

